# Structural and Social Disadvantage and Post-Migration HIV Vulnerability among African, Caribbean, and Black Communities in Ontario

**DOI:** 10.64898/2026.11.03.26364040

**Authors:** Wangari Tharao, Thabani Nyoni, Elsie Amoako, Adrianna Perryman, Danilia Fombang, Amrita Daftary, Mary Ndungu, Sandra Bullock, Jenna MacNeily, Soraya Blot, Valerie Pierre-Pierre, Mona Loutfy, Rupert Kaul, Darrell Tan, Anita Rachlis, Curtis Cooper, Irving Salit, Henry Luyombya, Shannon Thomas Ryan, Liviana Calzavara, the MSAFIRI Study Team

## Abstract

**Background:** African, Caribbean, and Black (ACB) communities in Canada experience a disproportionate burden of HIV, and a considerable share of infections among ACB immigrants is acquired after arrival in Canada. Yet how ACB people themselves understand the structural circumstances surrounding post-migration acquisition remains underexamined. This study asked how intersecting structural and social disadvantages shape post-migration HIV vulnerability among ACB communities in Ontario.

**Methods:** We conducted an interpretive secondary thematic analysis of 44 in-depth interviews from the MSAFIRI Study, read through intersectionality and structural-vulnerability lenses. Interviews were coded inductively and deductively by two analysts, with consensus adjudication and community review of the thematic structure.

**Results:** Five structural conditions organized participants’ accounts: racism as a structural condition of settlement; immigration precarity and the work of settling alone; economic precarity, including insecure work, poverty, and unstable housing; ruptured kin networks and isolation under migration; and gendered violence and homophobia. These conditions compounded one another, so that disadvantage in one domain deepened vulnerability in another. **Conclusion.** Post-migration HIV vulnerability among ACB communities is produced by intersecting structural disadvantages encountered in the receiving country context. Equitable HIV prevention requires structural and community-led responses that address immigration precarity, economic and housing insecurity, gendered violence, homophobia, and anti-Black racism, alongside biomedical and individual-level prevention.

## Introduction

African, Caribbean, and Black (ACB) communities continue to experience a disproportionate burden of HIV in Canada, and particularly in Ontario (Mbuagbaw et al., 2026). Although ACB populations represent a relatively small share of the Canadian population, they account for a markedly higher proportion of HIV diagnoses (Statistics Canada, 2024; Public Health Agency of Canada, 2026). This disparity reflects longstanding inequities rooted in racism, economic marginalization, and exclusionary social policies, not any inherent feature of ACB communities (Odhiambo et al., 2022). A considerable share of these infections is acquired after migration. In Ontario, an estimated 20% to 30% of immigrants from sub-Saharan Africa and the Caribbean who are living with HIV are thought to have acquired HIV after arriving in Canada (Ontario HIV Treatment Network [OHTN], 2016), and the MSAFIRI Study has documented the relational and sexual circumstances of post-arrival acquisition among ACB immigrants (Tharao et al., 2025). This evidence challenges the assumption that HIV among ACB people is primarily imported from countries of origin and redirects attention toward the conditions ACB migrants encounter following settlement. Understanding those conditions is therefore central to any equitable HIV prevention response.

This reorientation toward the receiving country context is consistent with a growing international literature. European estimates of post-migration acquisition among African and Caribbean migrants range from 2% to 63%, depending on the population and the method of estimation (Fakoya et al., 2015; Alvarez-del Arco et al., 2017; Gosselin et al., 2020), and post-migration acquisition has been linked to immigration precarity, housing instability, economic marginalization, and obstacles to healthcare access. North American evidence similarly implicates institutional and social contexts. Gendered patterns of service provision, including the limited representation and engagement of heterosexual ACB men within HIV prevention and support services, restrict access to HIV-related information, resources, and care (Baidoobonso et al., 2013). Vulnerability is thus shaped not only by individual circumstances but by the ways institutions and services are organized and made accessible to different groups within ACB communities.

Recognizing this, scholars have increasingly called for a departure from the behavioral frameworks that have long dominated HIV research and prevention. Such frameworks locate risk within individual knowledge, attitudes, and practices (J. K. Williams et al., 2009) and have too often encouraged deficit-oriented narratives that portray racialized communities as inherently vulnerable because of their behaviors, cultures, or beliefs. Structural models instead direct attention to the social systems within which risk is produced (Latkin et al., 2010), and C. C. Williams and colleagues (2009) argue that effective HIV prevention for Black women in Canada requires moving beyond risk-reduction models focused on individual behavior toward approaches that address the forces patterning health inequities. Canadian scholarship likewise emphasizes that racialized HIV disparities cannot be adequately understood without examining anti-Black racism, social exclusion, migration, and economic inequality (Baidoobonso et al., 2013; Nelson et al., 2019). A structural perspective thus shifts attention from what individuals do to the conditions under which choices become possible, constrained, or consequential.

For many ACB migrants, vulnerability emerges through settlement within institutions that were not designed around their social, cultural, or migratory experiences (Gagnon et al., 2022). Employment, housing, healthcare, education, and settlement systems are not encountered on equal terms; the intersection of race and migrant status shapes how individuals are received and supported (Gagnon et al., 2022; K. K. A. Williams et al., 2024). Labor-market exclusion, social isolation, and discrimination within institutions are, in this context, expressions of broader structural arrangements that position some groups differently from others within Canadian society (Odhiambo et al., 2022). Anti-Black racism is embedded within these social and institutional structures, shaping access to opportunities, resources, and support in ways that influence wellbeing and quality of life (Logie et al., 2013; Logie et al., 2016).

Immigration status and settlement experiences constitute additional structural determinants that profoundly shape health trajectories. Gagnon et al. (2022) describe immigration status as a foundational determinant of health because it structures access to legal protections and social services. For migrants with precarious status, uncertainty surrounding residency, work authorization, and eligibility for services can generate chronic insecurity and foreclose opportunities for stable settlement. Research among ACB immigrants living with HIV in Toronto shows how restrictive policies and legislative exclusions leave individuals medically uninsured, socially marginalized, and exposed to ongoing forms of institutional violence (Odhiambo et al., 2022, 2023). Economic insecurity and unstable housing frequently accompany these experiences, undermining wellbeing and heightening vulnerability long before HIV acquisition occurs.

Structural vulnerability is also produced through gendered and sexual power relations. Gender inequality, intimate partner violence (IPV), and homophobia can limit individuals’ capacity to negotiate safety, access support, or engage health systems without fear of stigma or harm (Fseifes & Etowa, 2023; Logie et al., 2011). Often experienced as traumatic, these forms of violence and exclusion generate psychological distress while erecting further barriers to healthcare engagement. Among ACB communities, they intersect with racism, migration experiences, and economic insecurity to create layered forms of disadvantage, and Canadian research has documented how these intersecting oppressions shape HIV vulnerability through unequal social relations (Etowa, Etowa, et al., 2022; Fseifes & Etowa, 2023).

To examine these processes, this study is informed by intersectionality and structural-vulnerability frameworks. Intersectionality holds that systems of oppression such as racism, sexism, heteronormativity, and immigration-related exclusion operate simultaneously and co-constitutively rather than as independent sources of disadvantage (Crenshaw, 1989, 1991). Within HIV research in Ontario, intersectionality has proven valuable for understanding how multiple forms of stigma and inequality shape the experiences of ACB communities (Logie et al., 2011). Complementing this perspective, structural-vulnerability and structural-violence frameworks direct attention to how social institutions, policies, and political-economic arrangements systematically produce unequal exposure to harm (Farmer et al., 2006; Quesada et al., 2011). Together, these frameworks locate HIV vulnerability within broader structures of power rather than within individual behavior alone.

Despite a growing body of literature on HIV among ACB communities in Canada, much of this scholarship has concentrated on HIV testing, prevention, risk behaviors, and individual-level interventions. Little is known about how ACB people themselves understand the structural circumstances surrounding HIV acquisition after migration. This study examined how intersecting structural and social disadvantages shaped post-migration HIV vulnerability among ACB people living with HIV in Ontario. The specific objectives were to: (1) describe the structural and social conditions that participants identified as surrounding their HIV acquisition after migration; (2) examine how these conditions intersected to shape vulnerability; and (3) identify implications for HIV prevention, practice, and policy. We drew on a secondary qualitative analysis of 44 interviews from the MSAFIRI Study and interpreted participants’ accounts through intersectional and structural-vulnerability lenses.

## Methods

### Design and Theoretical Orientation

This is a qualitative secondary analysis of the MSAFIRI Study, informed by the intersectional and structural-vulnerability frameworks outlined above (Crenshaw, 1989, 1991; Farmer et al., 2006; Quesada et al., 2011). Analytically, this meant treating racism, immigration policy, labor-market precarity, housing, and gendered power as the conditions under which ACB people came to be at risk, and reading these axes as co-constitutive rather than additive.

### Parent Study and Setting

MSAFIRI was a mixed-methods study of post-migration HIV acquisition among ACB communities in Ontario, comprising a cross-sectional survey (n = 108) and in-depth, semi-structured interviews with a subsample of 44 survey participants. Participants were recruited through purposive sampling from the Ontario HIV Treatment Network Cohort Study (OCS) across five HIV clinics in Toronto and Ottawa between January 2015 and March 2017. Interviews explored migration and settlement, relationships, health decision-making, and the circumstances surrounding HIV acquisition. The parent study’s design, eligibility criteria, and primary quantitative and qualitative findings are reported in full elsewhere (Tharao et al., 2025). The present paper draws on the interview data for a focused secondary analysis.

### Analytic Sample

The present analysis draws on all 44 interviews, focusing on extracts coded under Social Disadvantage, Traumatic Experiences, and Vulnerability. Consistent with the parent study (Tharao et al., 2025) and with conceptualizations of African, Caribbean, and Black communities as heterogeneous and diasporic rather than uniformly immigrant (Baidoobonso et al., 2013; Maheux & Do, 2019; Nelson et al., 2019), the sample comprises ACB people of Caribbean and sub-Saharan African origin together with a subset of Canadian-born participants of ACB parentage. Their inclusion is theoretically warranted: the structural conditions central to this analysis, namely anti-Black racism and social disadvantage, shape ACB lives across nativity and across generations (Baidoobonso et al., 2013; Jolly et al., 2013), such that vulnerability is patterned by racialized social location rather than by immigration status alone. Nativity is therefore treated as an analytic dimension rather than a screening criterion. Sample characteristics appear in Table 1.

**Table 1.** Sample Characteristics of the Analytic Sample (N = 44)

| **Characteristic** | **Category** | **n (%)** |
|---|---|---|
| Gender | Men | 23 (52.3) |
|  | Women | 21 (47.7) |
| Sexual orientation | Heterosexual (20 women, 7 men) | 27 (61.4) |
|  | Gay | 14 (31.8) |
|  | Bisexual or other | 3 (6.8) |
| Ancestry | Caribbean | 27 (61.4) |
|  | Sub-Saharan African | 17 (38.6) |
| Nativity and age at immigration | Immigrated at age 13 or older | 17 (38.6) |
|  | Immigrated before age 13 | 16 (36.4) |
|  | Born in Canada (ACB parentage) | 11 (25.0) |
| Mode of HIV acquisition | Heterosexual contact | 26 (59.1) |
|  | Sex between men | 16 (36.4) |
|  | Sexual contact with injection drug use | 2 (4.5) |
| Age at interview | Range (median) | 27–65 years (41) |
| Period of HIV acquisition | Range | Late 1980s–2016 |

*Note.* N = 44 African, Caribbean, and Black people living with HIV who completed in-depth interviews. Characteristics are drawn from the MSAFIRI qualitative interview tracking records. Bisexual and other sexual orientations, and the two acquisitions involving injection drug use (one heterosexual, one between men), are combined to protect confidentiality. Percentages may not sum to 100 because of rounding. ACB = African, Caribbean, and Black.

### Analytic Approach

The parent study used qualitative content analysis. For this paper, we conducted an interpretive thematic analysis through structural-vulnerability and intersectional lenses. TN and AP developed the codebook and coding structure for this analysis. AP and JM, both with qualitative data analysis experience, then independently coded the data using an inductive approach (several rounds of naïve coding) followed by a deductive approach (review for predefined categories based on the literature review and research objectives; Saldaña, 2016). No qualitative data analysis software was used. Coding was conducted manually in Microsoft Word, and the identified themes and illustrative quotes were then organized into a Microsoft Excel coding structure arranged by theme and sub-theme. The two coders resolved discrepancies through discussion to reach consensus; where consensus could not be reached, TN reviewed the coding and made the final decision. Auditing of the analytical processes and procedures was used to gain perspective and understanding and to enhance methodological rigour (Charmaz, 2006, 2014).

The coding process followed a stepwise approach. Using this codebook, the two coders began with an independent review of eight transcripts. Following a second independent review of these eight transcripts, each transcript was assessed for inter-rater agreement, prompting revisions to the codebook and coding structure. The eight transcripts were then reviewed and discussed further to adjudicate discrepancies until consensus was reached. Finally, the coders independently reviewed the remaining transcripts. From these codes, we (1) re-coded subthemes (e.g., anti-Black racism, immigration precarity, financial and job insecurity, unstable housing, and ruptured kin networks) into higher-order structural domains; (2) constructed themes that link individual accounts to the systems producing them; and (3) situated the themes within the structural-vulnerability and intersectional frameworks. Salience was gauged by the recurrence of each theme across participants’ accounts, and the resulting themes, sub-themes, and illustrative quotations are presented in Table 2.

**Table 2.**
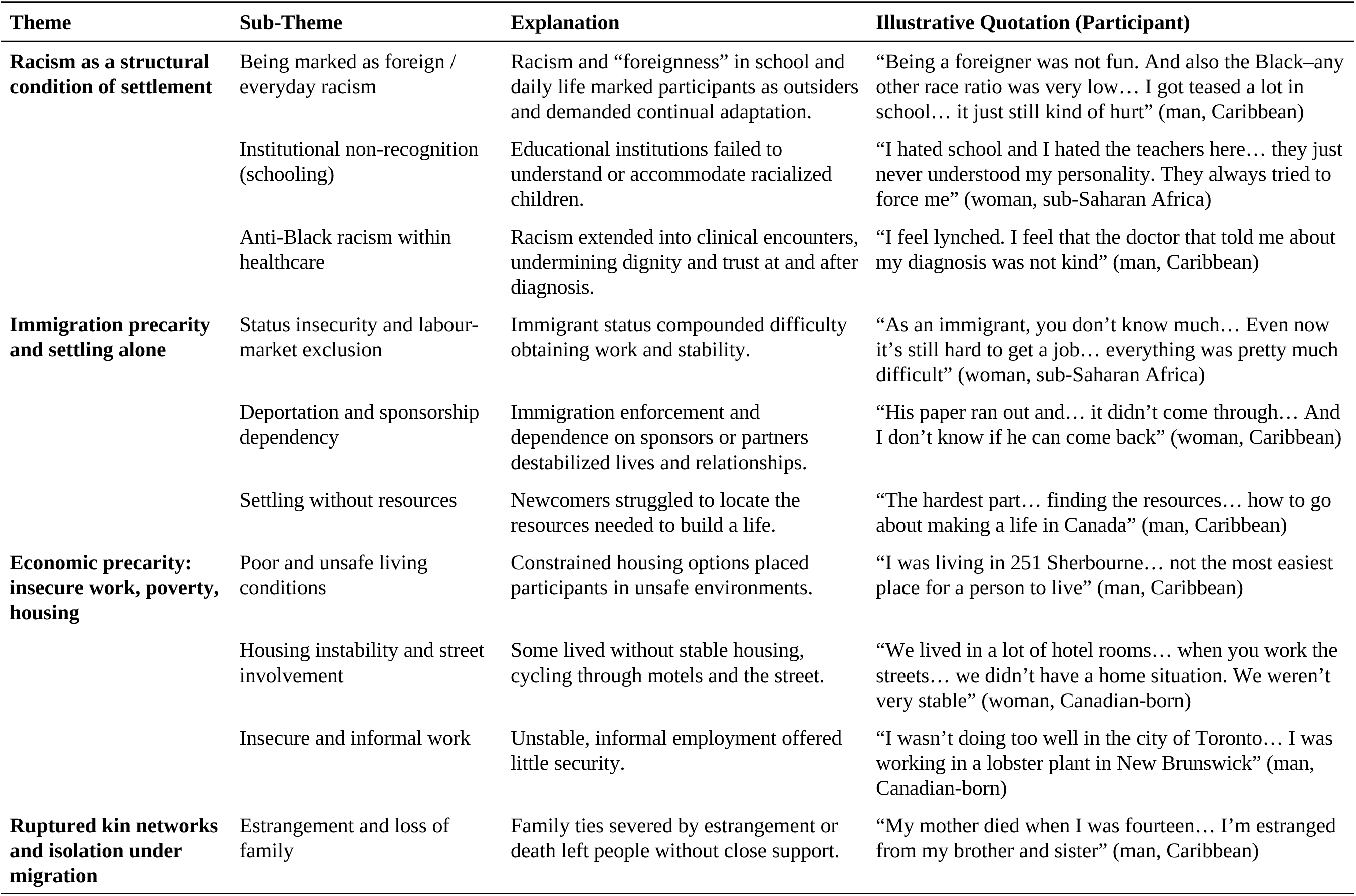
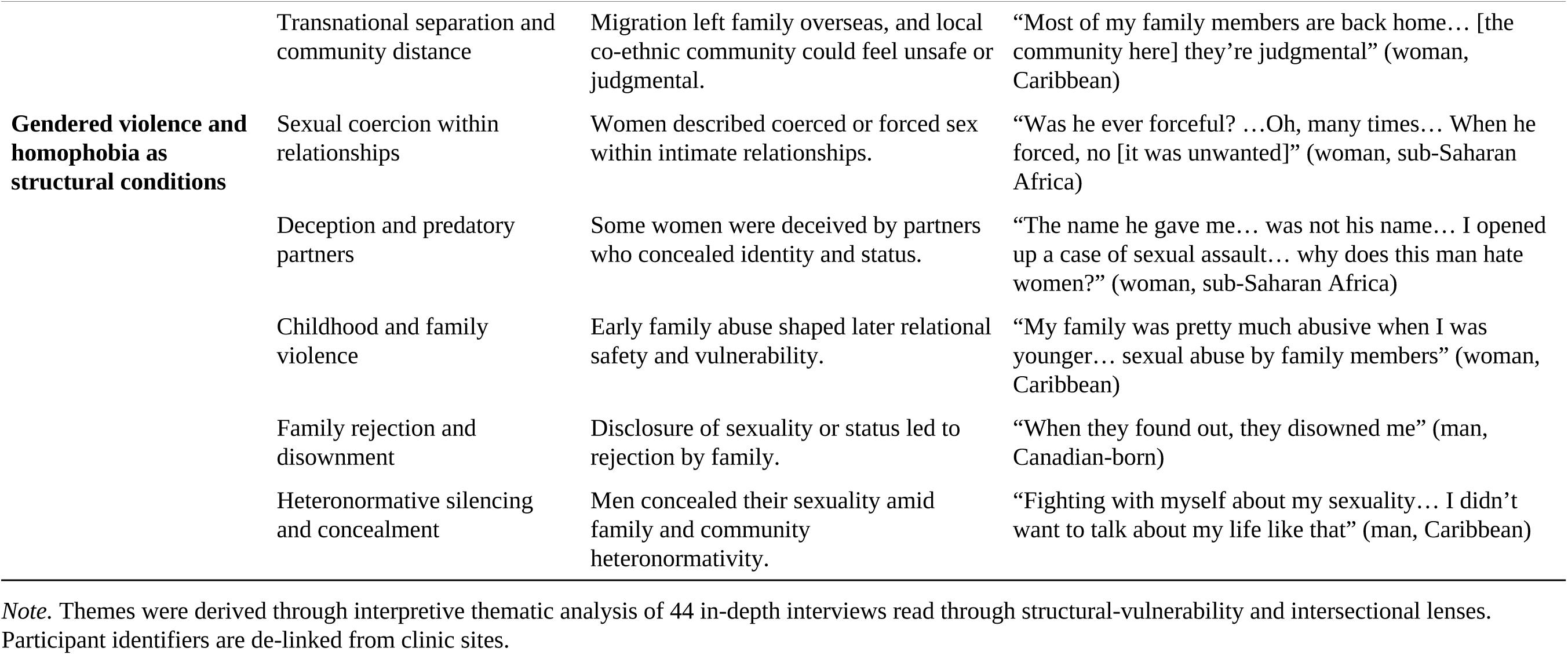
Structural Themes, Sub-Themes, and Illustrative Quotations.

### Reflexivity and Rigor

TN and AP, who developed the codebook, identify as Black, and JM, who coded the transcripts with AP, is a settler of European heritage. The analysis was conducted in partnership with Women’s Health in Women’s Hands Community Health Centre and the study’s Community Advisory Committee. Committee members reviewed the thematic structure to confirm that it reflected community experiences and to identify interpretations that could pathologize ACB communities. Trustworthiness was pursued using the criteria of Lincoln and Guba (1985): credibility (team-based interpretation and community review), dependability and confirmability (an audit trail from coded extracts to themes), and transferability (thick description of context). We retained accounts of agency and resilience alongside accounts of constraint to avoid a deficit narrative.

### Ethics

MSAFIRI received approval from the research ethics boards of the participating institutions, and all participants provided informed consent. Quotations are reported with participant identifiers de-linked from clinic sites, and small identifying details are generalized.

## Results

### Sample Characteristics

The analytic sample comprised the 44 ACB people living with HIV who completed in-depth interviews (Table 1). Twenty-three were men and 21 were women. Twenty-seven identified as heterosexual (20 women and seven men), 14 as gay men, and three as bisexual or another orientation. Twenty-seven participants were of Caribbean ancestry and 17 of sub-Saharan African ancestry. Seventeen had immigrated to Canada at age 13 or older, 16 had immigrated before age 13, and 11 were born in Canada to ACB parents. Most acquired HIV through heterosexual contact (n = 26), followed by sex between men (n = 16); two acquisitions involved injection drug use. Participants were aged 27 to 65 years at interview (median 41), and reported acquisition spanned the late 1980s to 2016. This diversity across gender, sexual orientation, region of origin, nativity, and life stage meant that the structural conditions described below were encountered from a range of social locations.

### Overview of Themes

Five structural themes organized participants’ accounts of the conditions surrounding post-migration HIV vulnerability: (1) racism as a structural condition of settlement; (2) immigration precarity and the work of settling alone; (3) economic precarity, including insecure work, poverty, and unstable housing; (4) ruptured kin networks and isolation under migration; and (5) gendered violence and homophobia as structural conditions of vulnerability. Table 2 presents the sub-themes and further illustrative quotations for each theme.

### Racism as a Structural Condition of Settlement

In participants’ narratives, anti-Black racism was not an isolated incident but a pervasive condition that shaped their lives from childhood or from arrival in Canada onward, influencing where they felt they belonged and how much effort daily life required of them. Several participants described racism encountered on arrival as something they had to absorb and adapt around. One woman who migrated from the Caribbean to Québec recalled: “*I was called ‘Chocolate’… I was teased a lot, especially in Québec. Black people had it really hard… I grew a thick skin to be able to stay*” (woman, Caribbean). Her account locates the harm in the receiving environment, in schools and among peers, and describes the “thick skin” she had to develop in order to stay. A Canadian-born participant of mixed ACB heritage described racism reaching back to early childhood: “*I dealt with a lot of racial tension… people were very, very prejudice when I was growing up… I remember asking my grandmother what colour my blood was*” (woman, Canadian-born). Her question to her grandmother shows how early she came to see her difference as a question of worth. This experience of racism began long before any sexual encounter and shaped the sense of self she brought to later health decisions. Together, these accounts position racism as a foundational condition that shaped settlement, belonging, and self-concept, and that operated through institutions and communities.

### Immigration Precarity and the Work of Settling Alone

Immigration status operated as a master condition that governed security, mobility, and access to basic supports, often leaving people to navigate unfamiliar systems without a map. Participants described status itself as a source of chronic uncertainty. One woman recalled: “*your immigration status, that’s if they accept you or no, you don’t know. You don’t know to be settled really here or no. You don’t know what is going on in your life*” (woman, sub-Saharan Africa). Her words render precarity as an ongoing state of suspension in which planning, including for health, is foreclosed by a decision held elsewhere. Others described settling with no institutional scaffolding at all. A man who arrived through Montréal recounted: “*I didn’t even know there was something like shelter. They didn’t tell me… somebody told me I have to go to welfare. I don’t know about welfare… So you had to figure everything out on your own*” (man, sub-Saharan Africa). The system into which he arrived furnished neither information nor support, so survival depended on chance encounters with strangers, conditions that crowd out preventive health engagement; he later noted that he saw a doctor “*only when I’m sick*.” Across these accounts, immigration precarity produced vulnerability by removing stability and withholding the information needed to reach the resources that make protective choices possible.

### Economic Precarity: Insecure Work, Poverty, and Unstable Housing

Insecure work and the poverty and housing instability that followed it repeatedly narrowed participants’ room to maneuver during the period surrounding acquisition. Job loss could cascade quickly into deeper precarity. One woman recounted: “*I found out that I was pregnant… so then I became jobless… I moved to her house and then from her house I went to live in a shelter*” (woman, sub-Saharan Africa). The sequence of pregnancy, job loss, dependence, and shelter shows how quickly economic insecurity stripped away options and how little buffer existed between employment and homelessness. A second participant described the churn of insecure work: “*I had been fired from probably the best job I ever had… I was going from job to job. At this point, I had no job*” (man, Canadian-born). His account conveys how the loss of stable employment unsettled the wider life context within which health and relationships were managed. These narratives show economic precarity constraining housing, stability, and bargaining power, so that what is conventionally termed “risk” was lived inside material constraints.

### Ruptured Kin Networks and Isolation under Migration

Migration frequently severed participants from the kin networks that elsewhere would have buffered hardship, leaving people to face settlement, illness, and risk with thinned support. The geographic scattering of family was a recurring structural feature. One woman stated plainly: “*Back home, it’s where my whole family is. In Canada, I have none*” (woman, sub-Saharan Africa). Her account locates isolation in the structure of migration itself: her family was absent by border, and that absence removed a first line of practical and emotional support at the very time settlement was hardest. For others, isolation compounded with the demands of survival work: “*I lost contact with my sister, which is the one I was closest to… we were workaholics… most times I get up, go to work, by the time I come home it’s very late… It’s like a machine*” (man, Caribbean). Here the erosion of close ties is bound up with labour conditions that consumed time and energy, so that the work required to survive in Canada corroded the relationships that might have sustained him. Across these accounts, ruptured kin networks emerge as a structurally produced exposure: migration and precarious work hollowed out the support systems through which people would otherwise have weathered risk.

### Gendered Violence and Homophobia as Structural Conditions of Vulnerability

Gendered power and heteronormativity operated as structural forces in participants’ accounts, surfacing as IPV and sexual coercion in women’s narratives and as homophobia and its transnational criminalization in the narratives of sexual-minority men. Both constrained the capacity to refuse, disclose, know, or seek help. For several women, exposure occurred through a partner’s coercion and concealment. One woman described a husband who knew his status and refused both protection and treatment: “*I asked him for several times to use a condom… he was asking by his doctor to take the medication. He didn’t take… That’s why I say he did it by purpose*” (woman, sub-Saharan Africa). Her account renders acquisition as the outcome of coercive control within marriage: she sought protection repeatedly and was overridden by a partner whose non-disclosure operated through a gendered imbalance of power.

For sexual-minority men, vulnerability was frequently structured by homophobia that long predated any encounter and reached across borders. One bisexual man described the conditions that pushed his sexuality and sexual health into silence: “*being Jamaican and saying that you’re into guys… was a huge problem so I just never [brought it up]… homosexuality in the south is banned and HIV and homosexuality go hand in hand, so all that stuff is just hush-hush and not brought to light*” (man, Caribbean). Here criminalization, communal silence, and the stigmatized coupling of HIV with same-sex desire foreclosed open identity and access to sexual-health knowledge. These legal, familial, and communal forms of homophobia shaped his risk. These accounts position gendered violence and homophobia as structural determinants on the same footing as racism, immigration precarity, and poverty: they governed who could negotiate safety and who was silenced into risk. Where participants voiced self-blame for these exposures, we read such statements as evidence of internalized structural narratives, and the analysis locates responsibility in the violence and the systems that enabled it.

These conditions did not operate in isolation. They compounded, as when a woman navigating immigration precarity and isolation also negotiated a coercive partner, or a man managing racism and economic insecurity was also silenced by homophobia, so that disadvantage in one domain intensified vulnerability in another. Participants were not passive before these conditions; they adapted, endured, and at times resisted. Yet their capacity to protect their health was shaped by the structures that surrounded them.

## Discussion

This study examined how intersecting structural and social disadvantages shape post-migration HIV vulnerability among ACB communities in Ontario. Across 44 in-depth interviews, participants located the conditions surrounding acquisition in racism, immigration precarity, economic precarity, ruptured kin networks, and gendered violence and homophobia. Estimates that 20% to 30% of HIV among sub-Saharan African and Caribbean immigrants in Ontario is acquired after arrival (OHTN, 2016), alongside European estimates ranging from 2% to 63% (Fakoya et al., 2015; Gosselin et al., 2020), establish that post-migration acquisition is substantial. The present findings specify the post-migration conditions through which that vulnerability is produced, extend the MSAFIRI Study’s account of the relational circumstances of post-arrival acquisition (Tharao et al., 2025), and ground those conditions in participants’ own accounts.

### The Structural and Intersecting Production of Vulnerability

The central contribution of this analysis is to show that post-migration HIV vulnerability among ACB participants was produced by structural conditions operating together. This patterning is best understood through intersectionality, which holds that racism, gendered power, heteronormativity, and immigration-related exclusion are co-constitutive (Crenshaw, 1989, 1991; Bowleg, 2012). The woman negotiating a coercive partner while insecure in her legal status, or the man managing economic precarity while silenced by homophobia, did not experience these conditions as separable exposures that could be summed; each shaped and intensified the others. Reading vulnerability in this way requires treating intersecting social positions, not single determinants, as the unit of analysis (Bauer, 2014). Structural-violence and structural-vulnerability frameworks complement this reading by locating harm in institutions, policies, and political-economic arrangements (Farmer et al., 2006; Quesada et al., 2011), and the findings are consistent with a syndemic understanding in which violence, economic hardship, and stigma cluster and interact to amplify health risk within contexts of structural inequity (Singer & Clair, 2003). Together, structural-violence and intersectionality theories explain unequal health burdens by directing attention to overlapping vulnerabilities and the upstream conditions that produce them (Snell-Rood et al., 2021). The compounding observed here is the mechanism these frameworks anticipate: disadvantage in one domain erodes the resources needed to withstand disadvantage in another.

These results align with and extend Ontario-based scholarship documenting how intersecting stigma, racial discrimination, and gender inequality shape the lives of ACB people living with HIV (Logie et al., 2011; Logie et al., 2013; Logie et al., 2016). That work has focused largely on the consequences of intersecting stigma for wellbeing after diagnosis. The present analysis examines the period surrounding acquisition and shows that the same structures implicated in post-diagnosis marginalization were already shaping exposure before diagnosis. Anti-Black racism, in particular, operated as a pervasive condition that shaped settlement, belonging, and self-concept from childhood or from arrival in Canada, which supports calls to treat racism as a fundamental cause of health inequity (K. K. A. Williams et al., 2024).

### Post-Migration Vulnerability and the Receiving Context

By foregrounding conditions encountered after arrival, the findings reframe a persistent narrative that attributes HIV among ACB communities to importation from countries of origin. Participants’ accounts of status insecurity, labour-market exclusion, unstable housing, and severed kin networks describe a receiving country context that generated vulnerability. This interpretation converges with European evidence linking post-migration acquisition to immigration precarity, economic marginalization, and barriers to care (Fakoya et al., 2015; Alvarez-del Arco et al., 2017; Gosselin et al., 2020), and with Canadian research showing how legislative exclusion and precarious status produce structural barriers to HIV care for ACB immigrants (Gagnon et al., 2022; Odhiambo et al., 2022, 2023). Our contribution is to demonstrate that these same structural forces shape vulnerability upstream of diagnosis, during the process of settlement itself, and to do so from the standpoint of ACB people. The limited engagement of heterosexual ACB men with HIV services noted in prior Canadian work (Baidoobonso et al., 2013; Etowa, Etowa, et al., 2022; Nelson et al., 2019) is echoed in accounts of men who encountered systems that furnished neither information nor support, underscoring that access is structured well before any individual decision about testing or protection.

### Resisting Deficit Narratives: Agency within Constraint

A consistent analytic commitment of this study was to resist deficit-oriented readings that locate HIV risk in the behaviours, cultures, or beliefs of racialized communities. Participants grew “a thick skin,” pieced together survival without institutional support, and at times named and resisted the injustices they faced. A structural lens reads such agency as exercised within, and constrained by, arrangements that distributed the capacity to protect one’s health unequally. This orientation is consistent with community priorities and with structural approaches that call for prevention to move beyond individual risk reduction toward the conditions that pattern health inequities (Latkin et al., 2010; C. C. Williams et al., 2009; Bowleg, 2012; Etowa, Tharao, et al., 2022). Recognizing both agency and constraint is central to a structural-vulnerability analysis.

### Implications for Prevention, Practice, and Policy

If vulnerability is produced structurally, then prevention must be structural as well. Biomedical tools such as pre-exposure prophylaxis and treatment as prevention remain essential, but their reach depends on the very conditions participants lacked: stable status, secure housing, economic footing, trustworthy services, and freedom from violence. Several implications follow. First, immigration status should be treated as a modifiable determinant of HIV vulnerability, with attention to expanding healthcare eligibility and reducing the insecurity generated by precarious status (Gagnon et al., 2022; Odhiambo et al., 2022, 2023). Second, responses to economic and housing precarity, including access to stable housing and income support during settlement, are HIV-prevention measures insofar as they restore the material ground on which protective choices become possible. Third, services must address gendered violence and homophobia directly, through trauma-informed and IPV-responsive care for ACB women and through affirming, confidential services for sexual-minority ACB men whose exposure was structured by criminalization and communal silence (Fseifes & Etowa, 2023; Logie et al., 2011). Fourth, the limited engagement of heterosexual ACB men warrants service models designed around, rather than incidentally accessible to, this group (Baidoobonso et al., 2013; Etowa, Etowa, et al., 2022). Across all of these, anti-Black racism within healthcare must be named and remediated so that clinical encounters build rather than erode trust (K. K. A. Williams et al., 2024). Such measures are most credible and effective when they are community-led and grounded in the priorities of ACB communities themselves (Etowa, Tharao, et al., 2022; Nelson et al., 2019), consistent with the community-partnered orientation of the present study.

## Limitations

This study has several limitations. As a secondary analysis, it was bounded by data generated for the parent study’s purposes; interviews were not designed to elicit every structural domain examined here, and questions tailored to this analytic focus might have yielded further detail. The interviews were conducted between 2015 and 2017, and aspects of the immigration and service context have since changed, although the structural conditions described remain salient. As a qualitative study grounded in the accounts of 44 ACB people living with HIV in two Ontario cities, the analysis prioritizes transferability and conceptual insight rather than statistical generalizability. Finally, the analytic sample includes Canadian-born participants of ACB parentage, retained for the reasons set out in the Methods. Their experiences of “post-migration” vulnerability differ from those of foreign-born participants, and the sample was not large enough to analyze nativity strata in depth; findings should therefore be read as speaking to ACB diasporic communities broadly rather than to foreign-born migrants alone.

## Conclusion

Post-migration HIV vulnerability among African, Caribbean, and Black communities in Ontario is the cumulative, intersecting outcome of structural conditions encountered in the receiving context. Racism, immigration precarity, economic and housing insecurity, ruptured kin networks, and gendered violence and homophobia converged to distribute the capacity to protect one’s health unequally, well before acquisition occurred. Equitable HIV prevention therefore requires structural and community-led responses that address these conditions directly, alongside biomedical tools, and that challenge the deficit narratives participants’ lives so clearly refute. Centering the standpoint of ACB people living with HIV offers not only a more accurate account of how vulnerability is produced but also a firmer foundation for prevention and health-system responses grounded in health equity.

## Declarations

### Ethics Approval and Consent to Participate

The MSAFIRI Study received approval from the research ethics boards of the participating institutions, including the Hospital Research Ethics Board (REB# 14-138), Sunnybrook Research Ethics Board (REB# 196-2014), Toronto General Hospital Research Ethics Board (REB# 14-7686-BE; University Health Network CAPCR ID 14-7686), University of Toronto Umbrella Ethics Board (Protocol Reference #29586), and St. Michael’s Research Ethics Board (OHSN-REB #20150802-01H). The Maple Leaf Medical Clinic was covered under the University of Toronto ethics approval (Protocol Reference #29586). All participants provided informed consent. Quotations are reported with participant identifiers de-linked from clinic sites, and small identifying details are generalized.

### Author Contributions

WT, LC, SB, RK, STR, VP-P, AD, HL, ML, DT, AR, IS, and CC co-designed the MSAFIRI mixed-methods study and developed the data collection processes. ML, DT, AR, IS, RK, and CC recruited participants through the participating clinics. SB, LC, and AD conducted the surveys and qualitative interviews, including quantitative analysis and qualitative coding. WT, STR, and VP-P reviewed the analysis and provided feedback.

TN conceptualized the current study, developed the codebook and coding structure with AP, made the final decision on unresolved coding discrepancies, interpreted the findings, and led manuscript drafting and revision. AP developed the codebook and coding structure with TN and independently coded the interview transcripts. JM independently coded the interview transcripts with AP, contributed to the resolution of coding discrepancies, and reviewed and approved the final manuscript. DF contributed to manuscript writing, review, editing, and formatting. WT provided guidance on study conceptualization, critical review, and editorial support, and coordinated feedback from co-authors. SB, AD, WT, and LC supplied information and contextual knowledge from the parent MSAFIRI study to support manuscript development. MN, EA, SBl, SB, VP-P, ML, RK, DT, AR, CC, IS, HL, STR, LC, and AP contributed to the interpretation of findings and critically reviewed the manuscript.

All authors read and approved the final manuscript.

### Corresponding Author

Correspondence to Thabani Nyoni, School of Social Work, Dalhousie University, Halifax, NS, Canada.

### Availability of Data and Materials

The data are available from the corresponding author upon reasonable request.

## Data Availability

All data produced in the present study are available upon reasonable request to the corresponding author.

## Acknowledgements

The MSAFIRI Study Team*, the Women’s Health in Women’s Hands Community Health Centre, and the Dalla Lana School of Public Health, University of Toronto, extend sincere gratitude to Dr. Frederic Crouzat and his team at Maple Leaf Medical Clinic, as well as collaborators at St. Michael’s Hospital, Sunnybrook Health Sciences Centre, Toronto General Hospital, and Ottawa Hospital Research Institute. We also thank the 108 participants across the five HIV clinics whose experiences made this study possible. We acknowledge the ongoing collaboration between the Dalla Lana School of Public Health and the participating institutions.

We further acknowledge the Community Advisory Committee, including WHIWH Community Health Centre, The African and Caribbean Council on HIV/AIDS in Ontario, The Black Coalition for AIDS Prevention, Africans in Partnership Against AIDS, AIDS Committee of Ottawa, Positive Living Niagara (formerly AIDS Niagara), People to People Canada, and Regional HIV/AIDS Connection (formerly AIDS Committee of London), for their support, guidance, and contributions to this study.

Finally, we gratefully acknowledge the late Prof. Robert S. Remis for his pivotal role in conceptualizing and initiating the MSAFIRI study and for his leadership in securing resources that supported its implementation.

*MSAFIRI Study Team: Keresa Arnold, Caroline Godbout, James Duncan Iveniuk, Amy Lee, Juan Liu.

